# Who carries the cost of interruptions? A qualitative study of doctors in UK hospital outpatient departments

**DOI:** 10.64898/2026.10.11.26365196

**Authors:** Shaun Boustani, Chase Staras, Antonia Rich, Ann Griffin

## Abstract

**Background:** Hospital outpatient departments (OPDs) deliver specialist care at scale, under close scrutiny over waiting times. Interruptions are common in this setting and are linked to error. Little is known about how they cause harm, for whom, and under what conditions.

**Methods:** Qualitative study using focus groups and interviews with 42 doctors working in UK National Health Service OPDs (19 sessions, September to November 2025). Participants were purposively sampled by clinical experience and self-reported patient complexity. Data were analysed using the Framework Method, informed by Cognitive Load Theory and Social Identity Theory, with independent double coding of 10% of transcripts and participant validation.

**Results:** We developed four themes. (1) *Interruptions as a structural and increasingly ambiguous feature of clinical work*. Interruptions were a normal part of clinical coordination, but new communication channels lacked shared norms about urgency, so each doctor had to judge what mattered. (2) *The hidden cognitive cost of interruptions*. The main cost came from returning to the interrupted task. This cost grew with complexity and was only partly offset by experience. (3) *The social fabric of interruption burden*. Transient teams weakened the norms that regulate interrupting, and approachability and hierarchy concentrated the burden on particular doctors. (4) *The social amplification of cognitive burden*. Understaffing multiplied cognitive costs. Doctors used familiarity and track record to decide which requests deserved attention, so concerns from junior or unfamiliar colleagues risked being discounted.

**Conclusions:** Interruptions are structural, and their impact depends on cognitive and social conditions. Reducing interruption frequency alone may miss the main risk. Interventions should support task resumption, protect capacity at busy times, rebuild shared communication norms and take account of the receiver’s capacity when designing escalation.

**WHAT IS ALREADY KNOWN ON THIS TOPIC:** Interruptions are common in clinical work and are linked to error, and laboratory studies place much of their cost in resuming the interrupted task.

**WHAT THIS STUDY ADDS:** In UK hospital outpatient departments, doctors’ accounts were consistent with these known mechanisms and suggest two further ones. Unfamiliar teams add cognitive load because each request has to be judged on its content, and under pressure, concerns raised by junior or unfamiliar colleagues may be discounted by the person receiving them.

**HOW THIS STUDY MIGHT AFFECT RESEARCH, PRACTICE OR POLICY:** Interventions should support task resumption, set shared norms about urgency, protect doctors’ capacity at busy times and design escalation with the receiver’s capacity in mind.

## Introduction

Hospital outpatient departments (OPDs) provide specialist assessment, diagnostics and treatment for a wide range of potentially serious conditions. In 2024–25 the NHS in England delivered 146.1 million outpatient appointments,^1^ and OPDs carry key responsibility for national targets on waiting times and cancer care. Anything that degrades the efficiency or safety of the outpatient consultation therefore matters.

Work interruptions are unexpected suspensions of attention or activity on an ongoing task.^2^ Research indicates they are common in clinical work. Over 90% of consultations are interrupted in some primary care settings,^3,4^ and hospital doctors are interrupted several times an hour.^5,6^ In a UK survey of 465 secondary care doctors, 61.1% reported being interrupted often or very often during outpatient consultations, and interruption frequency was independently associated with higher mental workload.^7^ Interruptions are linked to errors and to tasks left unfinished.^6,8,9^ Laboratory studies show that much of their cost lies in resuming the interrupted task. Suspended goals fade from memory, so resuming takes longer and fails more often when interruptions are longer or more demanding, and even interruptions of a few seconds can increase errors.^10–12^

Outpatient research has mostly counted interruptions, and much of it comes from primary care. We know little about how interruptions cause harm in hospital outpatient work, or who carries the cost. Interventions have mainly tried to reduce how often interruptions happen. However, many interruptions are necessary. A ‘do not interrupt’ intervention for nurses’ medication rounds reduced some interruptions, but nurses found it cumbersome and fewer than half supported making it hospital policy.^13^ This leaves two questions. The first is cognitive. How do interruptions disrupt clinical reasoning in OPDs, and how does this vary with task complexity and clinical experience? The second is social. Does interruption burden fall evenly across doctors, and how do team relationships shape how interruptions are experienced and recovered from?

We used two theories to address these questions (extended rationale in online supplemental file 1). Cognitive Load Theory (CLT) holds that working memory, the limited capacity for holding and using information in the moment, is finite.^14^ Interruptions add extraneous load. They take up working memory and leave a recovery cost when the clinician returns to the task.^15^ CLT predicts that this cost rises with the intrinsic load of the interrupted task, which in outpatients can be indexed by patient complexity,^16^ and that experience gives partial protection through automated routines (schemas).^14^ CLT therefore helps explain how much an interruption costs.

CLT says little about who bears that cost. Social Identity Theory (SIT) proposes that people understand themselves and others through group memberships such as professional role, seniority and team, and that these memberships shape communication and how demands are shared out.^17,18^ In healthcare, professional group boundaries shape how staff work together.^19^ SIT therefore predicts that status, approachability and familiarity shape who interrupts whom and how a request is judged.^20,21^ How an interruption is delivered also matters. Respectful or abrupt delivery affects both mood and the ability to refocus.^22^ Team churn, rotating staff and blurred hierarchies are common in healthcare, so these social conditions are likely to be important.

To our knowledge, this qualitative study is the first to use CLT and SIT together to examine how interruptions affect doctors in UK OPDs, and who carries the cost. We asked:

1. what interruptions doctors experience, and how they affect the interrupted doctor and the interrupter
2. through what cognitive mechanisms interruptions impose costs, and how task complexity and experience modify these
3. how team stability, status, familiarity and approachability shape the distribution of interruption burden
4. how cognitive and social processes interact.

A companion paper reports how doctors manage interruptions, using the same dataset.^23^

## Methods

### Design

We conducted a qualitative study using focus groups and individual interviews. This approach suited the limited evidence on how interruptions are experienced in OPDs and our interest in mechanisms. Reporting follows the Consolidated Criteria for Reporting Qualitative Research (COREQ).^24^

### Participants and sampling

Participants were doctors working in NHS hospital OPDs in the UK. Respondents to an online screener survey, which formed part of a national survey of UK secondary care doctors,^7^ were stratified into six groups by clinical experience (trainee, 0–19 years post-qualification, or 20+ years) and self-reported patient complexity (high or low). CLT and prior research identify these two factors as moderating the cognitive impact of interruptions.^25,26^ Stratifying by patient complexity, in addition to recording specialty, allowed us to examine whether the demands of a clinician’s caseload shaped their experience of interruption, as CLT predicts. This purposive approach ensured the sample spanned seniority and case mix (sampling details in online supplemental file 2). Of 207 eligible respondents, 42 doctors took part across 19 sessions (13 focus groups and 6 interviews), representing a broad range of specialties (table 1). Recruitment continued until the sample was judged to hold sufficient information power for the study aims.^27^ No participant withdrew. All names are pseudonyms.

**Table 1.**
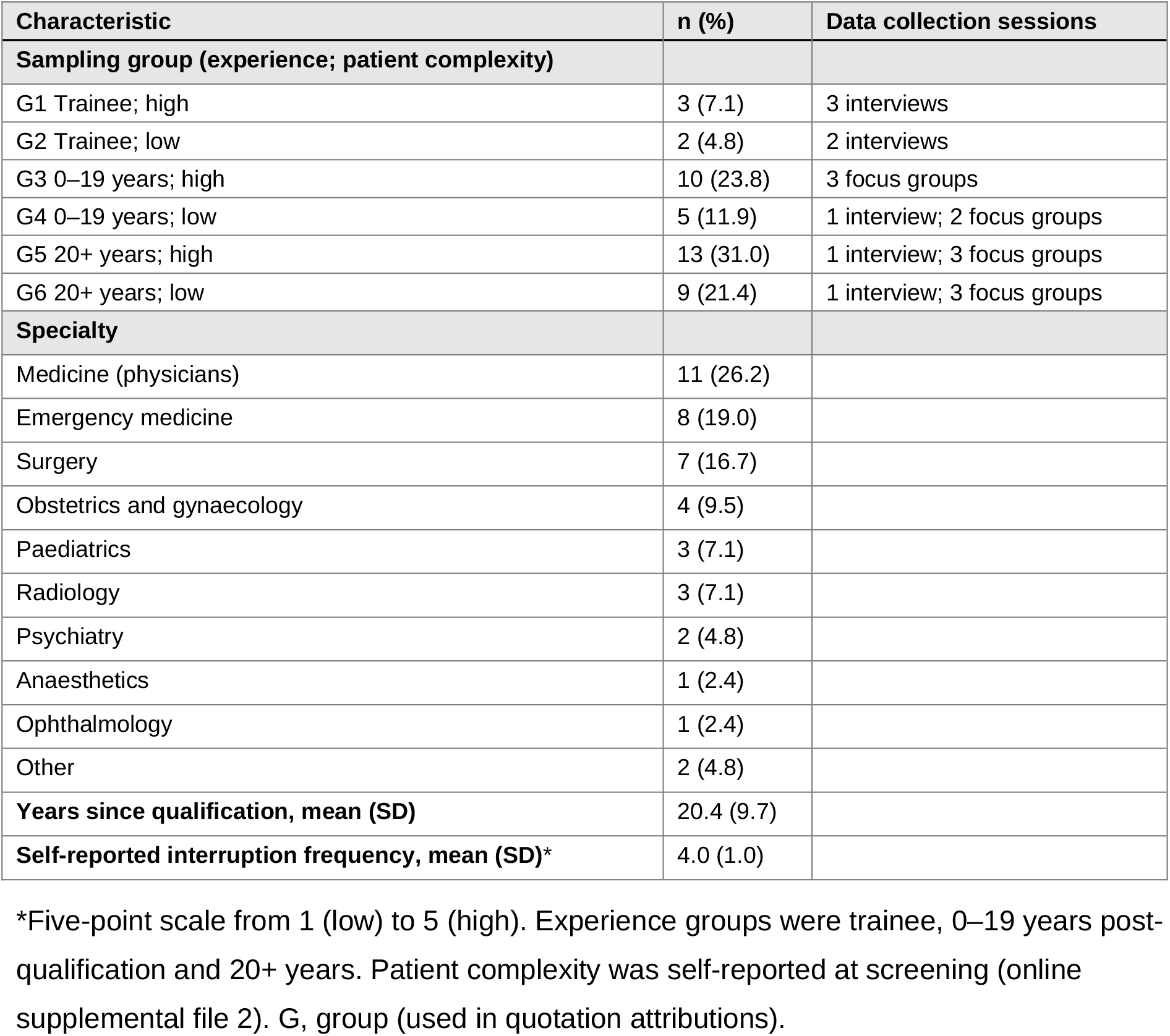
Participant characteristics (n=42)

| Characteristic | n (%) | Data collection sessions |
| --- | --- | --- |
| **Sampling group (experience; patient complexity)** |  |  |
| G1 Trainee; high | 3 (7.1) | 3 interviews |
| G2 Trainee; low | 2 (4.8) | 2 interviews |
| G3 0–19 years; high | 10 (23.8) | 3 focus groups |
| G4 0–19 years; low | 5 (11.9) | 1 interview; 2 focus groups |
| G5 20+ years; high | 13 (31.0) | 1 interview; 3 focus groups |
| G6 20+ years; low | 9 (21.4) | 1 interview; 3 focus groups |
| **Specialty** |  |  |
| Medicine (physicians) | 11 (26.2) |  |
| Emergency medicine | 8 (19.0) |  |
| Surgery | 7 (16.7) |  |
| Obstetrics and gynaecology | 4 (9.5) |  |
| Paediatrics | 3 (7.1) |  |
| Radiology | 3 (7.1) |  |
| Psychiatry | 2 (4.8) |  |
| Anaesthetics | 1 (2.4) |  |
| Ophthalmology | 1 (2.4) |  |
| Other | 2 (4.8) |  |
| **Years since qualification, mean (SD)** | 20.4 (9.7) |  |
| **Self-reported interruption frequency, mean (SD)\*** | 4.0 (1.0) |  |

\*Five-point scale from 1 (low) to 5 (high). Experience groups were trainee, 0–19 years post-qualification and 20+ years. Patient complexity was self-reported at screening (online supplemental file 2). G, group (used in quotation attributions).

### Data collection

Sessions were held online via Microsoft Teams between September and November 2025 and lasted approximately 55 minutes. Focus groups were grouped by experience level to limit hierarchical dynamics. They were led by one researcher and supported by a second, using a semi-structured topic guide covering interruption types, cognitive and social impacts, and management (online supplemental file 3). When only one participant signed up for a session, it was run as an interview by a single researcher to minimise any perceived power imbalance. Sessions were audio-recorded, auto-transcribed and checked against the recordings. Analytical memos were written during and immediately after each session.

### Analysis

Data were analysed using the Framework Method^28^ in NVivo 14, moving through familiarisation, coding, development and application of an analytical framework, charting and interpretation. Four researchers with backgrounds in cognitive psychology, social psychology, clinical medicine and health psychology developed a hierarchical codebook. It organised codes into five domains (nature of interruptions, cognitive factors, social dynamics, the intersection of cognitive and social factors, and management practices), and open coding was retained for new material. One researcher coded all transcripts and a second independently coded 10%, with high agreement (Cohen’s κ=0.97). Themes were developed from the coded data and interpreted through CLT and SIT. Draft findings were shared with all participants for validation.^29^ All 11 written responses agreed that the findings reflected their experience, and one added that interruptions compounded across a clinical session.

### Reflexivity

The facilitators’ backgrounds in cognitive and social psychology drew their attention to task switching and expertise, and to interpersonal and organisational dynamics, respectively. The wider team contributed clinical and health psychology perspectives. Team discussion kept both cognitive and social explanations in view throughout, and we actively looked for accounts that complicated emerging interpretations. Researchers had no prior relationship with participants.

### Patient and public involvement

Patients and the public were not involved in the design, conduct, reporting or dissemination of this research.

### Ethics

Ethical approval was granted by the UCL Research Ethics Committee on 20 May 2025 (ID 1132). Participants gave written consent via the screener survey and verbal consent to recording at the start of each session.

## Results

We developed four themes comprising ten subthemes (table 2). Quotations labelled Q1 to Q16 appear in table 2, and further quotations are in online supplemental file 4.

**Table 2.** Themes, subthemes and illustrative quotations.

| Theme and subtheme | Illustrative quotation |
| --- | --- |
| **Theme 1. Interruptions as a structural and increasingly ambiguous feature of clinical work** |  |
| 1.1 Structural normalisation of interruptions | **Q1** 'In the middle of conversations, phones are ringing, bleeps are going off, people are moving in between you and the person who's talking ... I couldn't put a number on it. They're countless. It's constant.' (Melissa, emergency medicine consultant, G5) |
|  | **Q2** 'If you are the leader, everyone will bother you ... No one is going to wait for me ... without these interruptions system cannot work ... You should be coping with these interruptions.' (Riha, paediatrics senior clinical fellow, G1) |
| 1.2 Collapse of urgency hierarchies across communication channels | **Q3** 'We've got phones, we've got bleeps, we've got electronic messaging services, we've got teams with, you know, emails ... there's not really an accepted standard of you know call me if it's an emergency. Text me if it's not |
|  | ... if I don't have all 7 methods of communication open at once. I'm going to miss something ...' (Catrina, paediatric oncology consultant, G3) |
|  | **Q4** 'This is my personal phone ... I don't understand why it's not an email like it's really not urgent.' (Daksha, geriatrician, G4) |
| **Theme 2. The hidden cognitive cost of interruptions** |  |
| 2.1 Parallel demands as the hidden cost of recovery | **Q5** 'Quite often [you'll] be sitting trying to write up some notes from a patient you've just seen while somebody is standing waiting to ask you about another patient and at the same time the phone rings ... And this goes on all day and I think, certainly cognitively, it's awful.' (Denise, acute medicine consultant, G5) |
|  | **Q6** 'Even brief interruptions break the cognitive flow ... But then there's the hidden cost of the interruption, the recovery time, and the cognitive residue ... that increases the risk of omissions and creates confusion trying to reconstruct the clinical narrative of the next patient.' (Darren, obstetrics and gynaecology, G4) |
| 2.2 Complexity as a multiplier | **Q7** 'A nurse grabbed me to say she's worried about a different patient ... it took me a minute to even remember what I was doing before ... It wasn't just a question. I'd had basically a whole new interaction and assessment ... compared to, say, any ECG that you can sign off in a couple of seconds. The more complex the interruption the, the harder it is to get back.' (Katrina, emergency medicine senior registrar, G2) |
| 2.3 Clinical experience as a partial buffer | **Q8** 'As we age, we lose our cognitive flexibility. I think I am aware I'm losing my flexibility ... we probably still work terribly more efficiently because we're more experienced and we're just going through algorithms and pattern recognising ... but they get to an age where they recognise ... this is more challenging for me.' (Alisha, respiratory consultant, G5) |
| **Theme 3. The social fabric of interruption burden** |  |
| 3.1 Transient teams and the erosion of shared norms | **Q9** 'There's the constant churn of agency staff. Even if you inducted everyone within an inch of their life ... you could create a wonderful system, but you'd still have the incoming agency.' (Steve, nephrology consultant, G3) |
| 3.2 Approachability as a mechanism of unequal burden | **Q10** 'I actually think that being approachable is a problem because there are 10 non-approachable people and if you want approachable, everyone will latch into you and they will potentially abuse your kindness ... it is sometimes a challenge to ask people for protected time.' (Macy, consultant, G3)* |
|  | **Q11** 'I actually think the balance of being somewhat unpopular and not as accommodating is something I'd rather have than be in a coroner's court explaining why I missed a result because me saying I was interrupted repeatedly is not a defence in that type of setting.' (Martha, respiratory consultant, G5) |
| 3.3 Status, conduct and the social regulation of interruptions | **Q12** 'If somebody comes in, is like I'm really, really sorry, is this a good time? ... You will have a much more open approach to them than somebody who just barges in and just starts talking.' (Max, emergency medicine consultant, G6) |
|  | **Q13** 'If it's the senior, sometimes even without prioritising, you have to do what they want you to do because it's the mental pressure you perceive by seniors ... I'll do it even if it's maybe more inconvenient than another request.' (Katrina, emergency medicine senior registrar, G2) |
| **Theme 4. The social amplification of cognitive burden** |  |
| 4.1 Systemic pressures as cognitive multipliers | **Q14** 'If that very same clinical question had been at the beginning of the shift it would have taken no mental effort ... but at the end of the shift it became a 10-minute problem because of everything that had happened beforehand.' (Victor, emergency medicine consultant, G3) |
| 4.2 The limits of familiarity and trust as heuristic filters | **Q15** 'If an F1 brings up a request for a full body MRI scan, you probably have the initial assumption of that's not the right thing to be doing ... Whereas if a consultant you've worked with for 20 years ... asks a question, your opening position is they probably have something sensible here. And even if I can't see |
|  | it immediately, it's worth my energy to find out what. So if somebody I know and respect and trust, there's a heuristic there.' (Owen, clinical oncologist, G6) |
|  | **Q16** 'It depends also on the track record of the colleague ... It's not if you like them or not. It's if they have a track record of previously high-quality interruptions or low-quality interruptions ... high-quality would be something that was time critical and contained the information that you needed.' (Megan, urology consultant, G6) |

\*Specialty not stated in transcript. Ellipses (...) indicate omitted text, and square brackets indicate words added for clarity. G, sampling group (table 1). F1, foundation year 1 doctor.

### Theme 1. Interruptions as a structural and increasingly ambiguous feature of clinical work

This theme describes how interruptions were experienced as an unavoidable part of secondary care, and how the growth of communication channels has made them harder to interpret.

#### Structural normalisation of interruptions

Participants described interruptions as a constant, structural part of secondary care. They came from colleagues, patients, phones, emails and electronic health record alerts, often several times within a single task (‘countless … constant’, Q1). Many participants were resigned to this (‘it’s just part of the job’; Steve, nephrology consultant, G3). For others, coping had become a professional standard and a marker of competence and leadership (Q2). Normalisation placed the burden of adaptation on individuals and hid the structural conditions that produce interruptions, which helped keep those conditions in place.

Participants were also clear that many interruptions are clinically necessary. The expectation to absorb them had developed when communication was simpler.

#### Collapse of urgency hierarchies across communication channels

Participants described a change in the kind of judgement interruptions now demand. They linked this to the growth of communication channels, including phones, bleeps, messaging services, Teams and email, without shared norms about urgency or etiquette (Q3). A request that once came as a knock on the door now arrived as a notification with no cue about priority. Urgent matters could be buried, and non-urgent requests could arrive at safety-critical moments. Clinicians sorted requests across channels in real time without reliable signals. One estimated that in ‘20 to 30%’ of cases requesters overstated urgency (Ryan, radiology registrar, G2), and others received non-urgent requests on their personal phones (Q4). Shared norms used to support this judgement. Each doctor now made it alone, before dealing with the interruption itself, and the normalisation described above meant they tended to absorb whatever arrived.

### Theme 2. The hidden cognitive cost of interruptions

Participants described an interruption as a layered event. Being interrupted, responding and returning each added to the cost, and the costs built up over a session in ways participants felt eroded confidence in their own clinical judgement.

#### Parallel demands as the hidden cost of recovery

Participants described holding a half-finished clinical picture in working memory while dealing with new, unrelated demands, such as writing up one patient while a colleague waited to ask about another and the phone rang (Q5). This felt exhausting and error-prone, especially when the interruption itself required careful reasoning. As one consultant put it, ‘it requires a lot of energy to go from one task to get fully concentrated in the other query and then go back’ (Macy, consultant, G3). The cost continued after the interruption ended.

Participants were most emphatic about the cost of returning to the interrupted work, the ‘hidden cost’ of ‘recovery time’ and ‘cognitive residue’ that raised the risk of omissions (Q6). An interruption that seemed brief and resolved could leave a trace that carried into later tasks. Current systems do not record or attribute this risk.

#### Complexity as a multiplier

Recovery cost grew with the complexity of both the interrupted task and the interruption, and was greatest when both were complex. Keeping a complex patient’s tests and plans in mind meant an interruption could send a clinician ‘back to square one’ (Charlotte, paediatric consultant, G6). Simple requests, such as signing a form or an ECG, needed little reorientation. An interruption that required assessing another patient needed a full change of focus and made returning harder (Q7). The cost of an interruption therefore depended on how its demands combined with those of the task. Complex queries arriving mid-decision were described as both common in high-acuity work and the most costly, so risk may cluster at moments that are already vulnerable. This needs direct testing.

#### Clinical experience as a partial buffer

Experience gave partial protection. Senior clinicians described being ‘well rehearsed’ and drew on diagnostic scripts and pattern recognition to pick tasks back up efficiently (Darren, obstetrics and gynaecology, G4). This protection was incomplete and changed over a career. Some later-career clinicians described losing cognitive flexibility, which made switching harder even though experience helped them handle task content quickly (Q8). The accounts suggest two kinds of vulnerability. Early-career doctors may face higher recovery costs because their schemas are less developed, and later-career doctors may recover content efficiently but find switching harder. Most participants were consultants, so the early-career pattern rests on fewer accounts. Clinical systems and workforce planning rarely take account of either.

### Theme 3. The social fabric of interruption burden

The meaning and impact of interruptions depended on who initiated them, under what conditions, and with what expectation of response.

#### Transient teams and the erosion of shared norms

In stable teams, a shared understanding of colleagues’ roles and workload quietly regulated when and how people interrupted. Most participants, however, described teams that changed constantly through rotations, agency staff and cross-specialty cover. Shared norms could not form in these teams, and even thorough induction could not keep up (Q9).

Colleagues new to a department did not know ‘when is it appropriate and who to mention things to’ (Nate, rheumatology, G3). Their interruptions felt inconsiderate, and participants described persistent frustration and emotional strain as a result, which they attributed to unfamiliarity with the department rather than ill intent. With channel norms also gone (theme 1), regulating interruptions fell to individuals, and it did not fall on all of them equally.

#### Approachability as a mechanism of unequal burden

Participants valued approachability as essential to team communication and patient safety. Yet approachable doctors became informal hubs for queries and absorbed a disproportionate share of interruptions (Q10). This burden clustered among conscientious mid-career and senior clinicians known for expertise and responsiveness, whose clinical and supervisory demands were already high. Participants described a ‘trade-off’ between being approachable and having space to work out ‘what do I need to do now’ (Eimear, stroke consultant, G3). Responses to this trade-off varied. Some accepted being ‘somewhat unpopular’ to avoid missing a result, noting that being interrupted would be no defence in a coroner’s court (Q11). Others saw approachability as non-negotiable because ‘it’s been shown time and time again that this kind of thing … improves patient safety’ (Daksha, geriatrician, G4). The openness that kept communication flowing was also the main route by which burden built up.

#### Status, conduct and the social regulation of interruptions

How an interruption was delivered shaped how it was received. Apologetic, well-timed approaches were tolerated, while colleagues who ‘barge in’ caused irritation and met a less open response (Q12). Seniority also mattered. Despite talk of flatter hierarchies, junior and mid-career doctors felt pressure to deal with senior colleagues’ requests straight away, even when the timing was poor (Q13). This pressure came from internalised obligation, without any explicit instruction. Less experienced doctors, whose schemas for recovery are less developed, were the least able to absorb the cost of interruptions and also the least able to refuse or defer them. Across this theme, the social organisation of clinical work shaped interruption burden in three ways. It weakened shared norms, it made approachability costly, and it set out who could interrupt whom and with what expectation of compliance.

### Theme 4. The social amplification of cognitive burden

Participants’ accounts showed that cognitive and social processes were intertwined. Cognitive load depended on the task and also on who was interrupting, how well they were known and the wider staffing context.

#### Systemic pressures as cognitive multipliers

Understaffing, rising workload and constant multitasking multiplied the burden of interruptions by putting several high-stakes demands into the same moment. One example was a lone registrar managing a critical case, a deteriorating ward patient and a consultant’s follow-up request at once (Riha, paediatrics senior clinical fellow, G1; online supplemental file 4). The burden also built up over a shift, so the same question carried different weight depending on when it arrived (Q14). The cognitive impact of interruptions is therefore tied to the organisational conditions in which they occur. Efforts that target individual behaviour without changing these conditions are unlikely to be enough.

#### The limits of familiarity and trust as heuristic filters

Familiarity and trust helped clinicians decide where to spend limited attention. Requests from long-standing, trusted colleagues were given the benefit of the doubt and judged ‘worth my energy’ to understand, while requests from junior or unfamiliar colleagues started from scepticism (Q15). Familiarity also worked through track record. Colleagues whose interruptions had proved time-critical and complete were seen as ‘high-quality’ interrupters, and others as low-yield (Q16). These tacit rankings helped clinicians cope with limited capacity. They also carried a risk. A request from someone without a track record had to be judged on its content, which took effort. Participants’ accounts suggest that when that effort was not available, the source of a request could decide how it was treated. Concerns raised by unfamiliar, junior or less articulate colleagues therefore risked being undervalued or delayed. Shortcuts that protected cognitive space created new risks.

## Discussion

### Principal findings

Doctors described interruptions in OPDs as constant, often necessary and increasingly hard to interpret. Their accounts were consistent with mechanisms known from laboratory and other clinical settings. The main cost of an interruption came from returning to the interrupted task, this cost grew with complexity, and experience only partly protected against it. The study also suggests two mechanisms that are less well described. Social conditions can create cognitive load, and the escalation of clinical concern can fail at the receiver.

Approachability and hierarchy concentrated the burden on particular doctors.

### Known mechanisms in a new setting

Laboratory research places much of the cost of interruption in resuming the suspended task,^10–12^ and observational studies link interruptions in hospital medicine to higher workload, errors and unfinished tasks.^5,6,9^ Our participants described the same pattern in outpatient consultations. Their accounts linked recovery cost to omissions, to complexity and to the build-up of load across a session. A national survey of outpatient doctors found that difficulty resuming interrupted tasks was associated with worse self-rated performance.^7^ Experience helped, as CLT’s account of schema automation predicts,^14^ but it did not remove the cost.

Participants also stressed that many interruptions are necessary, and attempts to reduce interruptions in other settings have had mixed acceptability.^13^ Interventions that aim only to reduce frequency may therefore miss the main source of risk. The safety value of an interruption lies in the information it carries, and the risk lies in returning to the task it disrupted. Recovery costs leave no record, so incident reporting is unlikely to capture them.

### Social conditions can create cognitive load

CLT describes how much an interruption costs but treats load as a property of the task. SIT describes how status, familiarity and norms distribute demands but says little about their cognitive cost. Our participants’ accounts link the two. With a familiar colleague, a doctor can use that colleague’s track record to judge a request quickly. Without that history, as in a transient team, the doctor has to judge each request on its content, and this appraisal uses working memory before the clinical question has even been addressed. Social conditions therefore help determine how much load an interruption creates, as well as who receives it. This fits evidence that professional group boundaries shape how healthcare staff interact,^19^ and suggests that team stability is a cognitive resource for clinicians. The loss of channel norms adds to this work. Digital communication systems have had unintended consequences in inpatient care,^30^ and our participants described them removing the cues that once signalled urgency, leaving more appraisal for each doctor to do alone.

### A second failure point at the receiver

Escalating a clinical concern depends on a speaker raising it and a receiver acting on it. Most work on speaking up has focused on the speaker, through psychological safety, graded assertiveness and structured communication tools.^31^ Receivers are known to respond differently depending on the speaker’s seniority and professional identity.^32–34^ Our findings suggest when this is most likely. Judging a request on its content takes capacity. When capacity is limited, people tend to rely on cues such as source credibility,^35,36^ and clinicians under cognitive load rely more on stereotypes and pattern recognition.^37–39^ A doctor who values junior colleagues’ concerns may still discount them when overloaded. Speaker-side interventions could therefore succeed on their own terms while concerns are still lost at the receiver, and protecting receivers’ capacity at busy times may matter as much as encouraging speakers. We present this as a proposition drawn from participants’ accounts, which our design cannot demonstrate. It predicts that discounting of concerns raised by junior or unfamiliar colleagues will increase with the receiver’s concurrent load, independent of the content of the concern.

### Who carries the burden

The burden of interruption was uneven. Approachability is set out as a core professional value,^40^ yet it drew interruptions towards the most collegial doctors, who often had the heaviest clinical and supervisory load. Hierarchy shaped who could say ‘not now’, and junior doctors, who are least equipped to absorb the cost, were least able to defer. Individuals also remained accountable for errors, as one consultant’s reference to the coroner’s court made clear (Q11). Discouraging approachability would harm safety culture, and asking individuals to set firmer boundaries would move the problem without solving it. The cost of approachability needs to be recognised by organisations. Both patterns are also related.

Being approachable, and judging requests by who sends them, are rational individual responses to conditions the individual did not create, and both generate harms of their own. Responses pitched at individual behaviour will therefore tend to relocate the problem rather than resolve it.

### Implications for practice and policy

Table 3 links each finding to possible responses. Five stand out.

**Table 3.** Implications of the findings for practice, policy and research.

| Finding | Implication | Possible responses |
| --- | --- | --- |
| The main cost of an interruption comes from returning to the interrupted task | Reducing frequency alone may miss the main source of risk | Structured hand-back and other task-resumption supports; evaluate effects on recovery costs and omissions as well as interruption rates |
| Recovery cost grows with the complexity of both the task and the interruption | High-complexity work needs protection | Protected time during complex consultations and peak demand |
| Communication channels have multiplied without shared urgency norms | Each doctor now judges urgency alone | Organisation-level agreements on which channel to use for which level of urgency; non-interrupting routes for routine requests |
| Transient teams erode the norms that regulate interrupting | Without a shared history, each request must be judged on its content, which adds load | Build interruption norms into team routines; consider team stability in rota design |
| Approachability concentrates burden on collegial, often senior clinicians | Approachability carries an unrecognised workload cost | Recognise interruption load and recovery time in job planning |
| Hierarchy limits junior doctors' ability to defer senior requests | Those least able to absorb the cost are least able to refuse | Senior clinicians to model and legitimise deferring non-urgent requests |
| Under load, the source of a request can decide how it is treated | Escalation of concern can fail at the receiver | Pair speaker-side escalation training with protection of receivers' capacity; test load-dependent discounting empirically |

1. Support task resumption, for example through structured hand-back, and evaluate interventions on recovery costs and omissions as well as interruption rates.
2. Agree shared urgency norms for each communication channel, and provide non-interrupting routes for routine requests.
3. Protect complex consultations and late-session work. Clinical decision-making can deteriorate over a session,^41^ and participants described the same interruption costing more late in a shift.
4. Recognise interruption load and recovery time in job plans.
5. Design escalation pathways with the receiver’s capacity in mind.

National policy is driving a shift from analogue to digital working in the NHS.^42^ Communication system design is a recognised contextual factor in safety,^43^ and without shared norms about how channels are used, more digital communication may add to interruption burden. In the national survey, individual strategy use was not consistently associated with lower workload,^7^ which supports system-level responses.

### Strengths and limitations

The study was designed around theory. Purposive sampling across experience and patient complexity, the two factors CLT predicts will shape cognitive impact, allowed comparison across groups. Combining a cognitive theory with a social theory produced explanations that neither theory offers alone and may be a useful template for interruption research. A multidisciplinary analytic team, double coding and participant validation strengthened credibility. The study also has limitations. All data were self-reported, and participants rated their own patient complexity. The sample came from UK NHS OPDs, which limits transferability to other health systems, although participants often drew on experience in ward and acute settings. Few trainees took part, probably because their duties are mainly ward-based, and most quotations came from consultants, so findings on early-career vulnerability need caution. We sampled only doctors. Interruptions involve whole teams, and research including nurses, allied health professionals and administrative staff is needed.

Future work should test whether task-resumption protocols, communication norms and more stable teams reduce recovery costs and improve safety, and should draw on the management strategies reported in a companion paper.^23^

## Conclusion

Interruptions in outpatient care are inevitable. Much of the harm they cause may be avoidable. Doctors’ accounts place the main risk in returning to the interrupted task, and show that social conditions shape how much load each interruption creates and who carries it. Responses should support task resumption, set shared norms for communication and protect doctors’ capacity at busy times. Because the adaptations doctors make are reasonable responses to conditions they cannot change, reducing frequency and relying on individual resilience will not be enough on their own.

## Supporting information

Supplementary Materials referenced in text

## Declarations

### Contributors

SB conceived and designed the study, obtained ethical approval and funding, led data collection, contributed to the analysis and drafted the manuscript. CS contributed to the conceptualisation of the study, co-facilitated data collection, led the coding and analysis of transcripts, and contributed to drafting the manuscript. AR contributed to the conceptualisation of the study, independently double coded a subset of transcripts, provided supervision and critically revised the manuscript. AG contributed to the conceptualisation of the study, provided supervision and critically revised the manuscript. All authors approved the final version of the manuscript. SB is the guarantor.

### Ethics approval

This study involved human participants and was approved by the UCL Research Ethics Committee on 20 May 2025 (ID 1132). All participants gave informed consent before taking part.

### Data availability statement

No data are available. Under the terms of the UCL research ethics approval (ID 1132), the transcripts and other participant data cannot be shared.

### Use of artificial intelligence

Claude (Anthropic), a generative AI tool, was used to help shorten and restructure the manuscript, format it to journal requirements and check the reference list. The authors reviewed and edited all output, verified all content and references, and take full responsibility for the manuscript.

## Acknowledgements

We thank the doctors who gave their time to take part in this study.

