## Supplementary Materials referenced in text for "Who carries the cost of interruptions? A qualitative study of doctors in UK hospital outpatient departments"

Online supplemental material

A qualitative study of the cognitive and social impact of interruptions on doctors in UK hospital outpatient departments

**Contents**

- Online supplemental file 1. Extended theoretical framework
- Online supplemental file 2. Sampling and participant validation
- Online supplemental file 3. Focus group and interview topic guide
- Online supplemental file 4. Additional illustrative quotations

### Online supplemental file 1. Extended theoretical framework

#### Cognitive Load Theory and the costs of interruption

Cognitive Load Theory (CLT) (Sweller et al., 2011) helps explain why interruptions are cognitively harmful and why the harm varies. CLT holds that working memory, the limited capacity for holding and using information in the moment, is finite, and that demands placed on it beyond the main task can degrade performance. Interruptions are a form of extraneous load. They are incidental to the clinical task, yet they take up working memory, break the flow of the task and leave a recovery cost when the clinician tries to return to it (Wirzberger et al., 2020). Recovery involves reactivating suspended goals, retrieving the partly completed task and re-establishing context. This is effortful and error-prone, and it is likely to be where most of the risk to clinical performance lies.

CLT also identifies conditions that change the size of this burden. Tasks with higher intrinsic load, such as those with more interacting elements, more uncertainty and harder decisions, leave fewer resources to absorb an interruption and make recovery more costly (Sweller et al., 2011). In secondary care outpatients, patient complexity is a meaningful index of this demand. Comorbidities, polypharmacy and psychosocial factors together raise the cognitive demands of the consultation (Nicolaus et al., 2022). Clinical experience is predicted to give partial protection. As practitioners develop automated routines and well-organised diagnostic schemas, familiar clinical material needs less effortful processing, which may reduce the cost of leaving a task and returning to it (Sweller et al., 2011). Whether this protection holds in the complex, high-demand environment of secondary care is an open question.

#### Social Identity Theory and the distribution of interruption burden

CLT accounts for the cognitive cost of interruptions but says little about who bears that cost, and why. Social Identity Theory (SIT) (Tajfel & Turner, 1979) offers a complementary lens. SIT holds that people understand themselves and others through social group memberships, such as professional role, seniority, specialty and team, and that these categories shape behaviour, communication and how interpersonal demands are shared out. Applied to clinical interruptions, SIT predicts that interruptions are patterned by the social structure of clinical work. This includes hierarchies of status and authority, norms of approachability and professional obligation, and the level of familiarity and trust within a team (Hogg & Terry, 2000; Janssens et al., 2025; Kerins et al., 2022).

The same interruption can therefore carry different costs in different social contexts (Janssens et al., 2025; Puranik et al., 2021). An interruption from a trusted colleague in a stable team with shared norms about when and how to interrupt imposes a different burden from one by an unfamiliar agency colleague in a transient team with no such norms. How an interruption is delivered also matters. A respectful or an abrupt approach affects both the clinician’s immediate mood and their ability to refocus. Research in organisational psychology shows that interpersonal interruptions can drain self-regulatory resources and also meet the need to belong, and that the relational quality of the exchange affects how disruptive the interruption proves (Puranik et al., 2021). Constant team churn, rotational staffing and blurred professional hierarchies are common in healthcare, so the conditions that SIT predicts will increase interruption burden, namely weak shared norms, uncertain status and low familiarity, are widespread.

### Online supplemental file 2. Sampling and participant validation

#### Screener survey and stratification

The screener formed part of a national online survey of UK secondary care doctors (Boustani et al., 2026). The survey invitation was emailed to the network of a UK medical defence organisation on 14 July 2025, with reminders on 28 July and 11 August, and the survey closed on 18 August 2025. Respondents could indicate interest in taking part in a focus group. The screener collected consent, role, specialty, years since qualification, self-reported interruption frequency (five-point scale from 1, low, to 5, high) and self-reported patient complexity.

Patient complexity was measured with a single item.

On a scale from 1 (low complexity) to 5 (high complexity), how would you rate the typical complexity of the patients you manage during consultations?

Response options 1 = Low, 2, 3, 4, 5 = High

Ratings were split at the median to give high and low complexity groups. In the full survey sample the median rating was 4 (n=415). Eligible respondents were then stratified into six groups by crossing experience level (trainee, 0–19 years post-qualification or 20+ years) with patient complexity (high or low). They were invited to sessions for their group so that focus groups were homogeneous for experience level.

#### Participant validation

A summary of draft findings was shared with all participants. Eleven written responses and two non-verbal affirmations (‘thumbs-up’ reactions) were received. Ten written responses agreed that the findings resonated with their experiences. The remaining respondent also agreed, and added that interruptions have a cumulative effect, building up across a clinical session. This exercise supported the credibility of the findings and refined the final outputs.

### Online supplemental file 3. Focus group and interview topic guide

*This semi-structured guide was used for online focus groups (grouped by similar levels of experience to mitigate power dynamics) and interviews. It is divided into sections exploring the nature of interruptions, their cognitive and social impacts, and strategies used for their management.*

**Opening and consent**

Hello, I’m [name] and I’ll be leading the interview/focus group today. Thank you for taking part. Before we begin I need to cover a few important ethical points.

Participation is voluntary and this session will take about 55 minutes. There aren’t any right or wrong answers and we’re just looking to get your opinions and experiences.

You’ve already given electronic consent via the screener; that consent covers your survey responses and the audio-recording for this focus group. If you would like to withdraw for any reason you can do so at any time during the session. Please note, however, that as transcripts will be pseudonymised, it will not be possible to remove contributions made to that point in the analysis.

Recordings and transcripts will be stored securely on [institution] systems, and only the research team will have access. We will also be taking notes during the session to remind us of important information.

If that’s all clear, I’ll start the recording. Do you have any questions or objections to being recorded?

**A. Introduction (5 minutes)**

- ‘As you know, we are interested in how unexpected interruptions, such as a colleague asking a quick question or a phone call in the middle of a consultation, affect your work. We also want to understand how these events might change the way you feel mentally, how they might influence interactions among team members, and how you manage an interruption once it’s occurred.’

**B. Experiences with interruptions (15 minutes)**

*Nature of interruptions*

- ‘To start, could you describe a typical interruption in your clinic? How often do these occur?’
- ‘What kinds of interruptions do you encounter most frequently (eg, trainee queries, phone calls, unexpected staff or patient interactions)?’

*Context and impact*

- ‘Are there any particular conditions (time of day, workload, clinic structure, staffing level) that make interruptions more or less easy to cope with?’
- ‘How do you think these interruptions influence your work?’
  - *Prompt* ‘And what about how they influence patient care and safety?’
- ‘What do you think makes an interruption particularly disruptive?’
  - *Prompt* ‘Is it the context, timing, who it comes from, or something else?’

**C. Cognitive impacts and task demands (15 minutes)**

‘I’d now like to ask some questions regarding the cognitive or mental impacts of interruptions. Sometimes we use the term “cognitive effort” or “cognitive load” to describe how much thinking or concentration is needed to complete a task. We’re interested in how interruptions may change this feeling.’

- ‘Thinking about a recent experience, did an interruption make it more challenging to get back on track with your consultation? If so, how?’
- ‘Are there certain parts of your work where interruptions seem to have a bigger impact on your concentration?’
  - *Prompt* ‘Does the complexity of patient information or the demands of the task influence your experience?’
- ‘What do you do to get back on track after an interruption? Are there any routines or steps you rely on?’
  - *Prompt* ‘What helps you decide whether to resume immediately, delay, or drop a task after being interrupted?’
- ‘Do you think these demands affect your well-being?’

**D. Social dynamics and interpersonal aspects (15 minutes)**

‘We’re also curious about how these interruptions influence your interactions with colleagues. By “team communication” we mean any conversation or exchange with other staff before, during or after a consultation.’

- ‘How do interruptions affect your interactions with colleagues?’
- ‘Do you think interruptions are perceived differently depending on whether they come from someone “inside” your team versus someone from another team or department?’
- If the interrupter is known, ask ‘Does your relationship with the interrupter affect the way you manage or respond to an interruption? What about their seniority?’
- ‘Have you observed any positive effects of interruptions (eg, brief, supportive interactions)? Under what circumstances?’
  - *Prompt* ‘Are there circumstances in which you don’t mind being interrupted?’
  - *Prompt* ‘Are there circumstances where being interrupted could be helpful?’
- ‘Now looking at some potentially negative impacts, can you share examples of when an interruption compromised team cohesion?’
  - *Prompt* ‘When interruptions go badly, what is usually happening in the background that contributes to this?’
  - *Prompt* ‘Do interruptions sometimes lead to clarifications or, conversely, to misunderstandings?’
- ‘Thinking about interruptions, both positive and negative, do they sometimes trigger emotions (eg, frustration, relief, reassurance) that shape how you respond?’

**E. Management and coping strategies (10 minutes)**

‘Finally, I’d like to discuss the ways you’ve learned to cope with interruptions once they’ve happened.’

- ‘What strategies have you or your team adopted to manage interruptions effectively?’
  - *Prompt* ‘In what situations do your strategies work well, and when do they not?’
  - *Prompt* ‘How do you know which tasks to prioritise? Can you give an example?’
  - If participants are struggling, prompt on the strategies identified in the survey, namely prevention, cognitive offloading and social strategies (putting people off).
- ‘Have you used any structured approaches (eg, handover or communication protocols) that you believe could work better?’
- ‘What barriers prevent the effective management of interruptions in your setting? What factors or resources would facilitate better management?’

**F. Future directions and close (5 minutes)**

- ‘Based on your experiences, what improvements or interventions would you suggest to reduce the negative impact of interruptions?’
- Thank participants and reiterate how their contributions will help improve both patient care and team well-being.

### Online supplemental file 4. Additional illustrative quotations

Quotations supplementing table 2 in the main manuscript. G, sampling group (see table 1 in the main manuscript). Ellipses (…) indicate omitted text.

| **Subtheme** | **Quotation** |
| --- | --- |
| 1.1 Structural normalisation of interruptions | ‘Your bleep goes off or you hear a noise that means you have to run … In the middle of conversations, phones are ringing, bleeps are going off, people are moving in between you and the person who’s talking so there’s audiovisual challenges … I couldn’t put a number on it. They’re countless. It’s constant.’ (Melissa, emergency medicine consultant, G5) |
|  | ‘It’s just part of the job.’ (Steve, nephrology consultant, G3) |
|  | ‘You cannot stop things happening around you. If you are the leader, everyone will bother you. Everyone will ask help from you. Everyone will continue to interrupt you. No one is going to wait for me. Otherwise, the system won’t work like that. You know, without these interruptions system cannot work … the ability of prioritising the things, the ability of to compose yourself, the ability of your response to these interruptions should be compact and different. You should be coping with these interruptions.’ (Riha, paediatrics senior clinical fellow, G1) |
| 1.2 Collapse of urgency hierarchies across communication channels | ‘20 to 30% of cases they’re overstating how urgent it is.’ (Ryan, radiology registrar, G2) |
| 2.1 Parallel demands as the hidden cost of recovery | ‘Because it requires a lot of energy to go from one task to get fully concentrated in the other query and then go back to fully concentrated on the previous task and there is clinical evidence, there’s scientific evidence that it will impact in the outcome.’ (Macy, consultant, G3)* |
| 2.2 Complexity as a multiplier | ‘When you’re actually trying to sort out something complicated, even if you’re doing lots of them, you’ve got to keep in mind that patient with all those tests, all this stuff, what the other person’s trying to do, do you need to change that? And I think that that, that’s … it’s too easy to be interrupted and you’re going back to square one again and it’s just hopeless.’ (Charlotte, paediatric consultant, G6) |
| 2.3 Clinical experience as a partial buffer | ‘I’m quite well rehearsed after all these years, and in what I say and the decision making in training takes a lot longer. Whereas I think it’s when you’re doing complex tasks, experience helps with that.’ (Darren, obstetrics and gynaecology, G4) |
| 3.1 Transient teams and the erosion of shared norms | ‘It can be frustrating if you’re the doctor on the clinic and you get informed [about a clinical case]. And sometimes it’s perhaps someone more new to the department … so it’s just that when is it appropriate and who to mention things to.’ (Nate, rheumatology, G3) |
| 3.2 Approachability as a mechanism of unequal burden | ‘It’s that trade-off isn’t it between wanting to be approachable and wanting to make them feel they can come and ask you, but that sort of like you say in your head working out the what do I need to do now.’ (Eimear, stroke consultant, G3) |
|  | ‘I think it’s really important when you’re working with a multidisciplinary team, when you’re perceived to be the leader … that people don’t have, they’re not afraid to come and talk to you. They do find you approachable. They do find you accessible … it’s been shown time and time again that this kind of thing increases and improves patient safety.’ (Daksha, geriatrician, G4) |
| 4.1 Systemic pressures as cognitive multipliers | ‘I was the only registrar working in the department … you are the only person dealing with so many juniors … you are dealing with a very sensitive and critical case … and someone interrupts in between. Oh, I need you because a patient in the ward is getting sick. Would you please come and deal with that. At the same time when I am leaving everything to deal with that patient and consultant just after asking if you have done this task which I allocated you earlier … it makes you mentally stressed … you need to prioritise.’ (Riha, paediatrics senior clinical fellow, G1) |

*Specialty not stated in transcript.
